# The Presence of Ambulatory Hypoxia as an Early Predictor of Moderate to Severe COVID-19 Disease

**DOI:** 10.1101/2020.12.14.20248209

**Authors:** Ajay Bhasin, Melissa Bregger, Mark Kluk, Peter Park, Joe Feinglass, Jeffrey Barsuk

## Abstract

**Background:** The importance of ambulatory hypoxia without resting hypoxia in COVID-19 is unknown. Ambulatory hypoxia without resting hypoxia may help objectively identify high-risk patients hospitalized with COVID-19. Interventions may be initiated earlier with sufficient lead-time between development of ambulatory hypoxia and other outcome measures.

**Methods:** We performed a retrospective study of adult patients hospitalized with COVID-19 from March 1, 2020 to October 30, 2020 in ten hospitals in an integrated academic medical system in the Chicagoland area. We analyzed patients who had daily ambulatory oximetry measurements, excluding patients who had first ambulatory oximetry measurements after the use of oxygen therapies (nasal cannula or advanced oxygen therapies). We determined the association of ambulatory hypoxia without resting hypoxia with the eventual need for nasal cannula or advanced oxygen therapies (defined as high flow nasal cannula, Bi-PAP, ventilator, or extracorporeal membrane oxygenation). We also calculated the time between development of ambulatory hypoxia and the need for oxygen therapies.

**Results:** Of 531 patients included in the study, 132 (24.9%) had ambulatory hypoxia. Presence of ambulatory hypoxia was strongly associated with subsequent use of nasal cannula (OR 4.8, 95% CI 2.8 – 8.4) and advanced oxygen therapy (IRR 7.7, 95% CI 3.4 – 17.5). Ambulatory hypoxia preceded nasal cannula use by a median 12.5 hours [IQR 3.25, 29.25] and advanced oxygenation therapies by 54 hours [IQR 25, 82].

**Conclusion:** Ambulatory hypoxia without resting hypoxia may serve as an early, non-invasive physiologic marker for the likelihood of developing moderate to severe COVID-19 and help clinicians triage patients and initiate earlier interventions.

## Introduction

Coronavirus-induced-disease-2019 (COVID-19) has caused a pandemic with 56,178,674 cases and 1,348,348 deaths worldwide.^1^ In our experience, we noted many inpatients with COVID-19 first exhibit hypoxia with exertion, then subsequently develop moderate to severe disease requiring nasal cannula or more advanced oxygen therapies. Early COVID-19 induces occult lung damage, noted by peripheral ground-glass opacities on imaging.^2^ We suspect patients at this stage exhibit ambulatory but not resting hypoxia. Several studies have characterized risk factors for severe COVID-19 based on patient-specific comorbidities and laboratory data. To our knowledge, no study has evaluated ambulatory hypoxia in absence of resting hypoxia as a risk factor for development of moderate to severe disease. Therefore, we aimed to evaluate if ambulatory hypoxia is associated with COVID-19 progression.

## Methods

We performed a retrospective observational study of adults (age ≥18 years) hospitalized with COVID-19 who had ambulatory oxygen measurements (without first developing resting hypoxia) at Northwestern Medicine (NM) between March 1, 2020 and October 30, 2020. NM is an integrated academic medical system in the Chicagoland area with 10 affiliated hospitals. We compared the associations between patients with and without ambulatory hypoxia and the use of nasal cannula (NC) or advanced oxygenation therapy [defined as high flow nasal cannula (HFNC), bi-level positive airway pressure (BiPAP), mechanical ventilation (vent) or extracorporeal membrane oxygenation (ECMO)]. We evaluated the time from the measurement of ambulatory hypoxia to initiation of NC and advanced oxygenation therapies. This study was approved by the Northwestern University Institutional Review Board.

### Procedure

We queried the Northwestern University Enterprise Data Warehouse (EDW) for all adult patients hospitalized with COVID-19 (ICD-10 code) who had ambulatory oxygen levels measured. The EDW is a complete database of clinical data extracted from the electronic medical records at NM. Hospital protocols within NM recommend that all patients with COVID-19 have once daily ambulatory oximetry screening, if able to participate. Any ambulatory oxygen saturation below 90% was considered hypoxia. Oxygen therapy outcomes included need for NC, HFNC, BiPAP, vent, and ECMO. Date and time of ambulatory hypoxia and initiation of all oxygen therapy outcomes were recorded. We *excluded* patients in whom oxygen therapy outcomes occurred before first ambulatory pulse oximetry measurement and those still hospitalized at the end of the study period.

Patient demographic and clinical data on admission including age, sex, race and ethnicity, body mass index (BMI), D-dimer, ferritin, and c-reactive protein (CRP) were recorded. The EDW query also included all ICD-10 codes for pre-existing chronic conditions. The Charlson comorbidity index for each patient was calculated based on admission ICD-10 codes to account for severity of illness at admission. For patients with multiple admissions, only data and outcomes during a chronologically first ‘index’ encounter were evaluated. Age, BMI, Charlson, D-dimer, CRP, and ferritin values were categorized for ease of interpretation. Time between ambulatory hypoxia and use of NC or advanced oxygen therapies was measured using the first occurrence of the oxygen therapy outcome.

### Analysis

Chi-square tests were used to evaluate bivariate associations between demographic and clinical variables and the proportion of patients with ambulatory hypoxia and subsequent use of nasal cannula or advanced oxygenation therapy. We used multiple logistic regression of the likelihood of nasal cannula to test the significance of ambulatory hypoxia because NC occurred in 13% of patients. We used multiple Poisson regression of the likelihood of advanced oxygen therapy to test the significance of ambulatory hypoxia because advanced oxygenation occurred in 6% of patients.^3 4^ We controlled for patients’ demographic and clinical characteristics including age, sex, race and ethnicity, BMI, and Charlson score, plus laboratory values that were significant at p<0.1 in bivariate associations. We performed sensitivity analyses substituting specific ICD-10 diagnoses including atrial fibrillation, coronary artery disease, congestive heart failure, diabetes mellitus, connective tissue disease, chronic obstructive pulmonary disease, hepatic failure with encephalopathy, human immunodeficiency virus, hypertension, leukemia or lymphoma, other immunodeficiencies, peripheral arterial disease, renal disease, solid tumor, and transplant in place of the Charlson score. R (3.6.1, Vienna, Austria) was used for analysis.

## Results

Our study included 531 patients with ambulatory oximetry measurements prior to oxygenation outcomes. Of these, 132 patients (24.9%) had ambulatory hypoxia. NC was required by 28.8% of patients with ambulatory hypoxia, compared to 8.0% of patients without it (p=2.4×10^−9^). Advanced oxygenation therapy was required by 18.9% of patients with ambulatory hypoxia, compared to 2.3% without it (p=4.6×10^−11^). Demographic and clinical variables are listed in Table 1.

**Table 1:** Percentage of Inpatients with COVID-19 with Each Demographics and Clinical Variable Who Underwent Ambulatory Oximetry and Developed Need For Oxygen Therapies.

| Characteristics | All Patients<br>(n=531)<br>%* | Nasal<br>Cannula<br>(n=70)<br>%* | Advanced<br>Oxygenation<br>Therapy<br>(n=34)<br>%* |
| --- | --- | --- | --- |
| Ambulatory Hypoxia Present<sup>a</sup> | 24.9 | 7.2 | 4.7 |
| Age (years)<sup>b</sup> |  |  |  |
| < 50 | 32.0 | 2.4 | 1.3 |
| 50-59 | 18.6 | 2.8 | 1.1 |
| 60-69 | 23.0 | 3.8 | 2.4 |
| ≥ 70 | 26.4 | 4.1 | 1.5 |
| Gender |  |  |  |
| Male | 50.1 | 6.2 | 4.0 |
| Female | 49.9 | 7.0 | 2.4 |
| Race and Ethnicity |  |  |  |
| Non-Hispanic White | 33.9 | 5.8 | 2.1 |
| Hispanic or Latino | 34.7 | 4.0 | 1.5 |
| Black or African American | 21.7 | 2.1 | 1.9 |
| Other Races and ethnicities | 9.8 | 1.3 | 0.9 |
| BMI (kg/m<sup>2</sup>)<sup>c</sup> |  |  |  |
| < 24.9 | 21.8 | 3.0 | 1.5 |
| 25.0 - 29.9 | 29.2 | 3.4 | 1.3 |
| 30.0 - 39.9 | 30.5 | 3.8 | 1.7 |
| ≥ 40 | 10.0 | 2.1 | 1.5 |
| Unmeasured | 8.5 | 0.9 | 0.4 |
| Admission D-Dimer (ng/mL) |  |  |  |
| 0-999 | 53.1 | 7.0 | 3.6 |
| ≥ 1000 | 5.8 | 0.8 | 0.8 |
| Unmeasured | 41.1 | 5.5 | 2.1 |
| Admission Ferritin<sup>d</sup> (ng/mL) |  |  |  |
| 0 – 499 | 44.8 | 4.9 | 2.4 |
| 500 – 999 | 13.2 | 1.9 | 0.8 |
| ≥ 1000 | 11.1 | 2.1 | 1.7 |
| Unmeasured | 30.9 | 4.3 | 1.5 |
| Admission CRP<sup>c</sup> (mg/L) |  |  |  |
| 0 – 99 | 55.6 | 6.8 | 3.2 |
| 100 – 199 | 13.4 | 2.6 | 1.3 |
| ≥ 200 | 4.5 | 0.9 | 0.8 |
| Unmeasured | 26.6 | 2.8 | 1.1 |
| Charlson Score<sup>e</sup> |  |  |  |
| 0-2 | 43.5 | 3.4 | 2.1 |
| 3-5 | 31.6 | 4.7 | 2.3 |
| ≥ 6 | 24.9 | 5.1 | 2.1 |
| Atrial Fibrillation | 10.0 | 1.9 | 0.8 |
| Coronary Artery Disease<sup>b</sup> | 4.1 | 1.1 | 0.4 |
| Congestive Heart Failure | 12.2 | 2.4 | 1.1 |
| Diabetes Mellitus | 32.4 | 5.5 | 2.6 |
| Connective Tissue Disorders | 2.8 | 0.4 | 0.2 |
| COPD | 8.9 | 1.5 | 0.8 |
| Liver Failure<sup>c,e</sup> | 1.1 | 0.6 | 0.4 |
| HIV | 0.8 | 0.0 | 0.0 |
| Hypertension | 58.8 | 8.3 | 4.3 |
| Leukemia or Lymphoma | 9.0 | 1.7 | 0.9 |
| Other Immunodeficiencies | 0.8 | 0.4 | 0.0 |
| Peripheral Artery Disease | 3.6 | 0.8 | 0.0 |
| Renal Disease<sup>b</sup> | 19.0 | 3.6 | 1.5 |
| Solid Tumor | 4.0 | 0.4 | 0.4 |
| Transplant<sup>b</sup> | 5.1 | 1.3 | 0.6 |
| <sup>a</sup>p < 0.0001 for nasal cannula and advanced oxygenation therapy<br><sup>b</sup>p < 0.1 for nasal cannula<br><sup>c</sup>p < 0.1 for advanced oxygenation therapy<br><sup>d</sup>p < 0.05 for advanced oxygenation therapy<br><sup>e</sup>p < 0.05 for nasal cannula<br>*All percent values are row percents relative to all patients (n=531) |  |  |  |

Multiple logistic and Poisson regression results confirmed that the presence of ambulatory hypoxia was strongly associated with the likelihood of using NC (OR 4.8, 95% CI 2.8 – 8.4) and advanced oxygenation (IRR 7.7, 95% CI 3.4 – 17.5); Table 2. Results were unchanged in the sensitivity analysis. Ambulatory hypoxia preceded NC use by a median 12.5 hours [IQR 3.25, 29.25] and advanced oxygenation therapies by 54 hours [IQR 25, 82].

**Table 2:** Multiple Logistic and Poisson Regression Results for the Association of Ambulatory Hypoxia with the likelihood of Nasal Cannula or Advanced Oxygenation use in 531 Inpatients with COVID-19.

| Characteristics | Nasal Cannula |  | Advanced Oxygenation |  |
| --- | --- | --- | --- | --- |
|  | OR (95% CI) | p | IRR (95% CI) | p |
| Ambulatory Hypoxia Present | 4.8 (2.8 - 8.4) | $3 \times 10^{-8}$ | 7.7 (3.4 - 17.5) | $8 \times 10^{-7}$ |
| Age (years) |  |  |  |  |
| < 50* | - | - | - | - |
| 50-59 | 1.5 (0.6 - 3.6) | 0.40 | 1.0 (0.3 - 3.2) | 0.97 |
| 60-69 | 1.2 (0.4 - 3.1) | 0.75 | 1.7 (0.5 - 6.2) | 0.40 |
| ≥ 70 | 0.6 (0.2 - 1.8) | 0.35 | 0.9 (0.2 - 4.1) | 0.88 |
| Gender |  |  |  |  |
| Male | 0.8 (0.5 - 1.4) | 0.47 | 1.6 (0.7 - 3.6) | 0.23 |
| Female* | - | - | - | - |
| Race and Ethnicity |  |  |  |  |
| Non-Hispanic White* | - | - | - | - |
| Hispanic or Latino | 0.8 (0.4 - 1.7) | 0.62 | 0.9 (0.3 - 2.5) | 0.85 |
| Black or African American | 0.4 (0.2 - 0.9) | 0.02 | 1.3 (0.5 - 3.4) | 0.59 |
| Other Races and ethnicities | 1.0 (0.4 - 2.6) | 0.98 | 2.1 (0.6 - 6.7) | 0.23 |
| BMI (kg/m2) |  |  |  |  |
| < 25.0* | - | - | - | - |
| 25.0 - 29.9 | 1.0 (0.5 - 2.2) | 0.95 | 0.7 (0.2 - 1.9) | 0.48 |
| 30.0 - 39.9 | 1.0 (0.5 - 2.3) | 0.91 | 0.8 (0.3 - 2.2) | 0.64 |
| ≥ 40 | 2.3 (0.9 - 6.2) | 0.10 | 2.6 (0.8 - 7.9) | 0.10 |
| Unmeasured | 1.1 (0.3 - 3.4) | 0.87 | 0.7 (0.1 - 3.5) | 0.65 |
| Admission CRP (mg/L) |  |  |  |  |
| < 100* | - | - | - | - |
| 100 – 199 | - | - | 1.9 (0.8 - 4.5) | 0.96 |
| ≥ 200 | - | - | 1.7 (0.7 - 4.1) | 0.81 |
| Unmeasured | - | - | 1.1 (0.4 - 2.9) | 0.68 |
| Admission Ferritin(ng/mL) |  |  |  |  |
| < 500* | - | - | - | - |
| 500 – 999 | - | - | 1.5 (0.5 - 4.7) | 0.94 |
| ≥ 1000 | - | - | 0.9 (0.3 - 2.6) | 0.28 |
| Unmeasured | - | - | 1.1 (0.3 - 3.5) | 0.77 |
| Charlson Score |  |  |  |  |
| < 3* | - | - | - | - |
| 3-5 | 2.4 (1.1 - 5.5) | 0.03 | 1.4 (0.5 - 3.9) | 0.55 |
| ≥ 6 | 4.6 (1.8 - 12.3) | 0.002 | 1.4 (0.4 - 5.1) | 0.63 |
| *Denotes reference category |  |  |  |  |

## Discussion

Our study shows that the presence of ambulatory hypoxia in absence of resting hypoxia is strongly associated with the subsequent development of moderate to severe COVID-19. In our patient population, ambulatory hypoxia occurred several hours before the need for NC and advanced oxygen therapies. This is important because it may enable clinicians to start therapeutic treatments such as remdesivir^5^ and/or dexamethasone^6^ earlier. It may also help health systems effectively identify patients most likely to require ICU-level care, which may improve hospital throughput especially when hospitals are near or at capacity.

Ambulatory oximetry has been used to help determine severity of cardiopulmonary diseases including heart failure,^7^ pulmonary hypertension,^8^ chronic obstructive pulmonary disease,^9^ and interstitial lung disease.^10^ To our knowledge, the presence of ambulatory hypoxia has never been shown to acutely predict cardiopulmonary disease progression. Recently, some authors have advocated for ambulatory oximetry measurements to evaluate patients for discharge readiness after resolution of COVID-19 symptoms,^11^ a strategy already adopted in our hospital. We present the first study showing that ambulatory hypoxia predicts worsening pulmonary disease in patients with COVID-19.

Our study has several limitations. First our study was performed at one health network with a relatively small number of patients potentially limiting generalizability. However, the associations between ambulatory hypoxia and worsening oxygenation were statistically very strong. Second, we excluded patients who did not have ambulatory oximetry measurements. Ambulatory oxygen measurement is part of the admission order protocols in our health system, so patients without measurements likely represented a sicker cohort unable to ambulate or those discharged quickly. Patients without measurement may also have been cared for by clinicians who did not order ambulatory oximetry, given lacking evidence. Third, we did not directly measure the presence of resting hypoxia because it was often not documented in the medical record before the use of oxygen therapy. The use of oxygen therapy was used as a surrogate for resting hypoxia. Finally, we excluded all patients who used oxygen therapy before ambulatory oximetry measures. These patients likely underwent ambulatory oximetry to determine discharge readiness. Alternatively, there may have been a small number of these patients who were given NC treatment for subjective comfort without resting hypoxia, who then developed ambulatory hypoxia.

Future randomized studies should be performed to evaluate earlier interventions in patients with COVID-19 and ambulatory hypoxia without resting hypoxia. An important extension of our findings could be the use of ambulatory oximetry measurement in triaging both outpatients and inpatients with COVID-19. Ambulatory hypoxia may help determine whether admission and inpatient monitoring with continuous pulse oximetry is warranted.

## Data Availability

The data is available is available for review upon reasonable request

## References

1. Dong E, Du H, Gardner L. An interactive web-based dashboard to track COVID-19 in real time. Lancet Infect Dis 2020;20(5):533–34. doi: 10.1016/S1473-3099(20)30120-1 [published Online First: 2020/02/23]

2. Wang Y, Dong C, Hu Y, et al. Temporal Changes of CT Findings in 90 Patients with COVID-19 Pneumonia: A Longitudinal Study. Radiology 2020;296(2):E55–E64. doi: 10.1148/radiol.2020200843 [published Online First: 2020/03/20]

3. Zhang J, Yu KF. What’s the relative risk? A method of correcting the odds ratio in cohort studies of common outcomes. JAMA 1998;280(19):1690–1. doi: 10.1001/jama.280.19.1690 [published Online First: 1998/12/01]

4. Zou G. A modified poisson regression approach to prospective studies with binary data. Am J Epidemiol 2004;159(7):702–6. doi: 10.1093/aje/kwh090 [published Online First: 2004/03/23]

5. Beigel JH, Tomashek KM, Dodd LE, et al. Remdesivir for the Treatment of Covid-19 - Final Report. N Engl J Med 2020;383(19):1813–26. doi: 10.1056/NEJMoa2007764 [published Online First: 2020/05/24]

6. Group RC, Horby P, Lim WS, et al. Dexamethasone in Hospitalized Patients with Covid-19 - Preliminary Report. N Engl J Med 2020 doi: 10.1056/NEJMoa2021436 [published Online First: 2020/07/18]

7. Gheorghiade M, Follath F, Ponikowski P, et al. Assessing and grading congestion in acute heart failure: a scientific statement from the acute heart failure committee of the heart failure association of the European Society of Cardiology and endorsed by the European Society of Intensive Care Medicine. Eur J Heart Fail 2010;12(5):423–33. doi: 10.1093/eurjhf/hfq045 [published Online First: 2010/04/01]

8. Rubin LJ. The 6-minute walk test in pulmonary arterial hypertension: how far is enough? Am J Respir Crit Care Med 2012;186(5):396–7. doi: 10.1164/rccm.201206-1137ED [published Online First: 2012/09/04]

9. Stoller JK, Panos RJ, Krachman S, et al. Oxygen therapy for patients with COPD: current evidence and the long-term oxygen treatment trial. Chest 2010;138(1):179–87. doi: 10.1378/chest.09-2555 [published Online First: 2010/07/08]

10. Khor YH, Goh NS, Glaspole I, et al. Exertional Desaturation and Prescription of Ambulatory Oxygen Therapy in Interstitial Lung Disease. Respir Care 2019;64(3):299–306. doi: 10.4187/respcare.06334 [published Online First: 2018/11/01]

11. Hussain T, Saman HT, Yousaf Z. Identification of Exertional Hypoxia and Its Implications in SARS-CoV-2 Pneumonia. Am J Trop Med Hyg 2020;103(4):1742–43. doi: 10.4269/ajtmh.20-1012 [published Online First: 2020/08/28]

